# Efficacy of acupuncture for chronic prostatitis/chronic pelvic pain syndromes on quality of life: study protocol for a randomized, sham acupuncture-controlled trial

**DOI:** 10.1101/2022.12.23.22283882

**Authors:** Yu-Long Ding, Huai-Yu Wang, Yuan Ji, Shuo Zhang, Peng-Fei Yuan, Hong-Chao Zhao, Yan Guo, Xiao-Di Xie

## Abstract

**Background:** Chronic prostatitis/chronic pelvic pain syndromes (CP/CPPS) is a common heterogeneous disease that seriously impacts patients’ quality of life (QoL). Acupuncture therapy has been widely used in China for various urinary diseases and symptoms, including chronic prostatitis. The results of several randomized controlled studies from different countries support that acupuncture can relieve the symptoms of CP/CPPS. Still, most randomized controlled trial (RCT) trials focus on symptom relief in patients, and the evidence on improving the QoL is insufficient. This study aims to assess the near-term and long-term efficacy of acupuncture in improving QoL in patients with CP/CPPS.

**Methods/Design:** This is a double-arm, parallel, participant-blinded RCT. 70 male CP/CPPS subjects aged 18-50 will be randomly allocated to either the acupuncture group or the sham acupuncture group. Participants will receive acupuncture or sham acupuncture treatment thrice a week over eight weeks for 24 sessions. The primary outcome will be the change in the total score of QoL compared with the baseline after eight weeks of treatment and 24 weeks of follow-up. The expectancy of acupuncture, blinding, and safety will also be assessed. A two-sided test will perform all statistical analyses, and a *p*-value of less than 0.05 will be considered statically significant.

**Discussion:** This study aims to provide quantitative clinical evidence of acupuncture effectiveness and safety in improving the QoL in patients with CP/CPPS.

**Clinical Trial Registration:** This trial is registered at chictr.org.cn (Identifier: ChiCTR2100051115)

## Introduction

CP/CPPS is a common genitourinary disease in the adult male population. This disease is mainly manifested by repeated and long-term pain, discomfort in the pelvic area, and different degrees of lower urinary tract symptoms. A substantial number of patients experience mental symptoms and abnormalities in sexual function, which adversely affect clinical efficacy and rehabilitation^[1]^.

According to the National Institutes of Health (NIH) classification, chronic CP/CPPS belongs to type III prostatitis. Type III prostatitis includes two subtypes: type IIIA (inflammatory CPPS) and type IIIB (non-inflammatory CP/CPPS). With the deepening of the study of CP/CPPS, experts realize that the etiology of CP/CPPS is complex, with different mechanisms, different clinical manifestations, different disease processes, and different responses to treatment of heterogeneous clinical syndrome, so curing CP/CPPS is not a realistic goal. At present, the clinical drug treatment of the disease is mainly symptomatic treatment, such as antibiotic treatment, beta-blockers, nonsteroidal anti-inflammatory drugs, anxiety medication and antidepressants, and botanical agents are widely used in clinical practice. However, the effect of the above treatment measures is not ideal, and there are specific adverse reactions and long-term use of patients with poor compliance.

In recent years, more and more attention has been paid to the QoL of patients. QoL is patients’ sense of approval and satisfaction compared to their expected or reachable functional state. It is a subjective feeling. In the clinical studies of different diseases, it is found that doctors often pay more attention to the significance of changes in objective indicators, and the understanding of the subjective state of patients is often inaccurate or not completely accurate, but the most important thing for patients is their subjective QoL. For example, the Atrial Fibrillation (AF) load, as a measurable and objective indicator, is highly valued by clinicians. However, whether it has a special significance for the patients is still being determined^[2]^. Clinical observation shows that some patients with persistent AF do not have any symptoms, while some patients with paroxysmal AF may experience severe symptoms such as dyspnea during the attack. The use of less toxic treatments than existing standard therapies to improve the QoL of patients with advanced non-small cell lung cancer (NSCLC) has not achieved the expected effect^[3]^. Therefore, from the point of view of patients and caregivers, it is more important to accurately assess the impact of the disease on the overall health of patients rather than just focusing on objective indicators. Acupuncture has been widely used in many urinary diseases and symptoms in China, including chronic prostatitis^[4-6]^. Several randomized controlled trials from different countries show that acupuncture can relieve the symptoms of CP/CPPS^[5-8]^. The results of a multicenter large sample RCT trial published in ANN INTERN MED in 2021 showed that 8-week acupuncture could effectively improve the symptoms of patients with moderate and severe chronic prostatitis / chronic pelvic floor pain syndrome (CP/CPPS). The clinical effect lasted at least half a year without involving patients’ QoL^[9]^. Most of the available data focus on symptom improvement in CP/CPPS patients, as controlling symptoms is an ideal way and way to improve QoL. However, symptoms and QoL cannot be equated, and the scores of QoL in patients with CP/CPPS may change in different dimensions because of their various symptoms. The evidence is insufficient on the efficacy of acupuncture studies in improving the QoL in patients with CP/CPPS. Currently, we regard acupuncture as an effective TCM rehabilitation method for treating CP/CPPS. Therefore, we assume that true acupuncture will have better clinical efficacy than sham acupuncture in improving QoL, symptom score, and long-term effectiveness.

## Methods and Analysis

### Study Design

A double-arm, parallel, participant-blinded randomized controlled trial (RCT) will be conducted at the Beijing Fengtai Hospital of Integrated Traditional Chinese and Western Medicine, a level A tertiary teaching hospital. The protocol has been registered in the China Clinical trial Registry (Project No. ChiCTR2100051115), and the study protocol has been approved by the Ethics Committee of Beijing Fengtai Hospital of Integrated Traditional Chinese and Western Medicine (Project No.: 19-Scientific Research-12-20). A total of 70 patients will be recruited, and all patients enrolled in the trial will voluntarily sign the informed consent form. Patients were randomly assigned to either the acupuncture treatment group (Acu Group) or the sham needle control group (Sham Group). Assessors will evaluate efficacy indicators before treatment, eight weeks after treatment, and 24 weeks after completion of therapy.

Beijing University of Chinese Medicine (BUCM) will be responsible for data management and statistics. The flowchart of the trial is shown in Figure 1, and Table 2 shows the schedule of the measurements.

**Figure 1.**
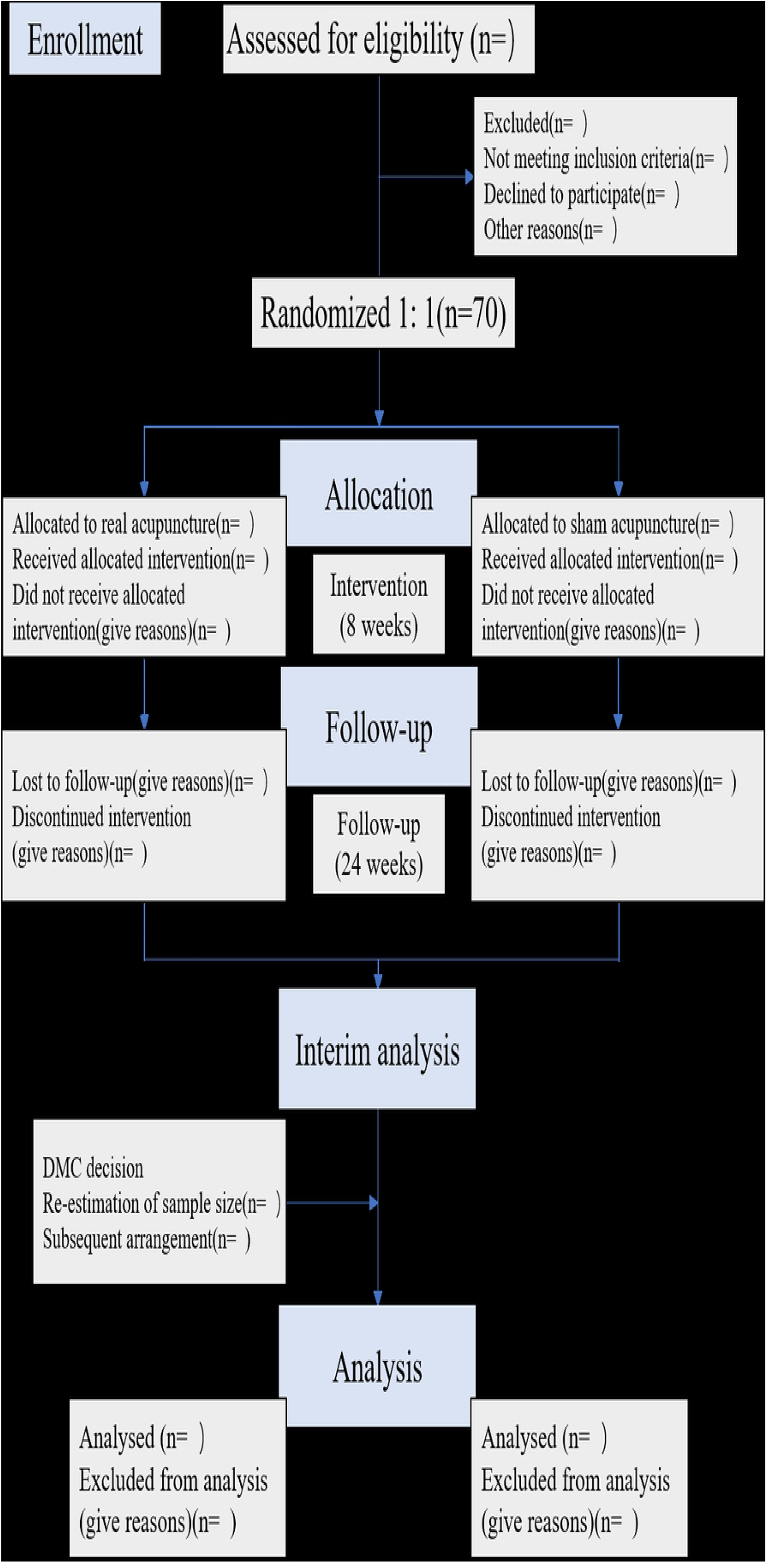
Flow chart of the trial. DMC, the data monitoring committee

**Figure 2.**
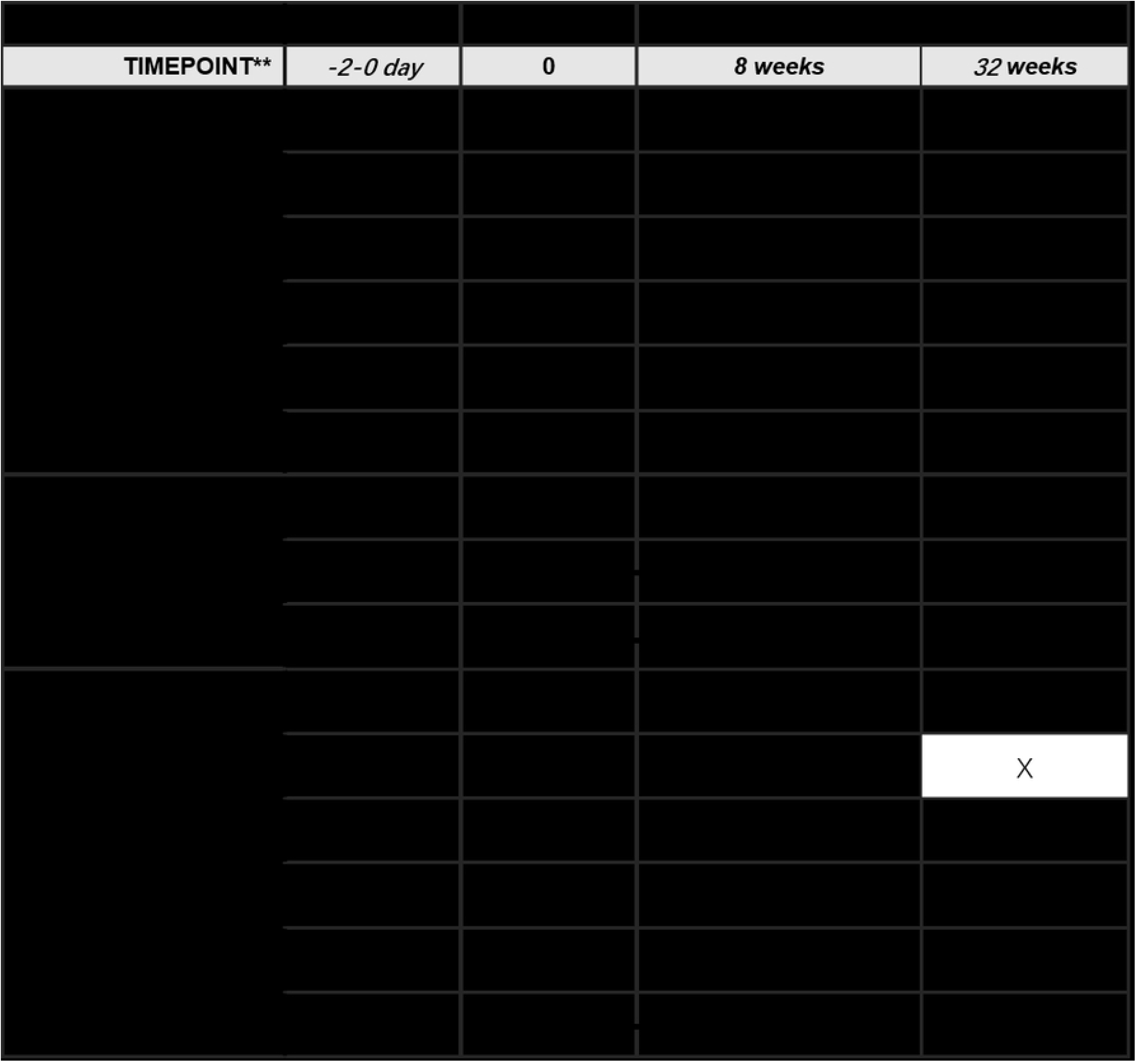
Study schedule showing the time points for enrollment and assessment ×, required; SF-36, The MOS item short-form health survey; IPSS, International Prostate Symptom Score; NIH-CPSI, The National Institute of Health Chronic Prostatitis Symptom Index; VAS, Visual analog scale.

### Participant Recruitment

This project is expected to recruit 70 male patients between January and December 2022. A urologist will screen CP/CPPS patients through medical history, physical examination, and laboratory examination. All participants will have to meet the CP/CPPS diagnostic criteria agreed upon by the NIH: the discomfort or pain of the pelvic area will last for at least three months in the past six months. The 2-glass test and other examinations, including prostatic fluid cultures, prostate-specific antigen evaluation, urine flow rate, and residual urine examination, will collect the prostatic fluid/urine specimen microbiology. Patients aged 18 to 50 years with National Institutes of Health Chronic Prostatitis Symptom Index (NIH-CPSI) total score ≥15 are eligible to participate. Participants will be screened if they agree to sign an informed consent form. Research assistants will conduct recruitment.

### Inclusion Criteria

The criteria for inclusion in this study are the following: (1) the clinical manifestations were repeated and persistent prostate pain accompanied by abnormal urination and mental and neurological symptoms in the last six months, and no infection or other obvious pathological conditions were confirmed. The symptoms lasted for more than three months. The main clinical manifestations are a pain in the surrounding tissue radiated by the prostate as the center. Joint pain, distension, or discomfort in the scrotum, testis, lower abdomen, perineum, lumbosacral, medial thigh, etc.: abnormal urination is characterized by frequent urination, urgency, pain, burning of the urethra, residual urine, or white discharge from the urethra when getting up in the morning, end of urine or defecation. Mental and neurological symptoms include dizziness and tinnitus, insomnia and dreaming, anxiety and depression, or even sexual dysfunction such as impotence and premature ejaculation. (2) aged 18 to 50 years; (3) NIH-CPSI total score ≥15; (4) Voluntarily participate in this experiment and sign the informed consent form.

#### Exclusion Criteria

Potential participants will be excluded for any of the following: (1) bladder outlet obstruction, bladder hyperactivity, neurogenic bladder, interstitial cystitis, cystitis glandularis, sexually transmitted diseases, cancer in situ and other bladder tumors, prostate cancer, urinary tract infection, other types of prostatitis, etc.; (2) residual urine ≥ 100ml by transabdominal ultrasonography, the maximum urinary flow rate is <15ml/s; (3) use of drugs to relieve the symptoms of CP/CPPS or receiving non-drug therapy for CP/CPPS (acupuncture, biofeedback, etc.) during the previous 1 month, and taking drugs that affect the function of the lower urinary tract; (4) Symptomatic urinary tract infection and urinary system organic diseases; (5) Diseases affecting the function of lower urethra, such as multiple sclerosis, Alzheimer’s disease, Parkinson’s disease, spinal cord injury, cauda equina injury, stroke and multiple system atrophy, etc.; (6) Patients with severe heart, lung, brain, liver, kidney and hematopoietic system diseases, mental illness and apparent cognitive impairment.

#### Randomization and Blinding

A randomized controlled design was adopted in this study. The random number of this study will be generated and sealed by the network department of our hospital and will be kept by the managers of the research group who do not participate in the project. Depending on the information received from the envelope, consecutive participants will be randomly assigned 1:1 to either the Acu group or the sham group. The acupuncturist will be given to perform acupuncture treatment for two groups. The study coordinator will not participate in the assessment or treatment. They will inform patients that they may receive either “the less painful acupuncture arm specifically developed for this study” (sham group) or the “traditional Chinese acupuncture arm” (Acu group). Efficacy assessors, acupuncture operators, and statisticians will be separated. Non-acupoint micro-acupuncture will allow the subjects to be blinded and evaluated in a blinded way after the last treatment.

### Interventions

Participants will receive acupuncture or sham acupuncture for 30 minutes in the supine position thrice a week for eight weeks. In addition, each treatment form and evaluation form will be completed and signed by patients and researchers. Two acupuncturists each will be responsible for the two groups. Before treatment, participants will be told they would receive traditional acupuncture with relatively deep acupuncture or minimal acupuncture with relatively shallow acupuncture at non-acupoints. Participants will receive treatment according to appointments to maintain blindness and avoid communication. The acupoints used are located in the lumbosacral region and the back of the tibia, which is difficult for participants to see when they lie prone. Participants may feel the needle penetrate the skin, but they are usually unable to judge the depth of the needle. To test the success of the blind method, within five minutes of two treatments in the eighth week, participants were asked if they had received traditional acupuncture in the previous weeks (yes, no, or unclear).

The use of drugs or other therapies for CP/CPPS will be avoided during the study unless participants’ symptoms are intolerable. Other medicines for CP/CPPS symptoms, such as α blockers, are not allowed in principle: all out-of-regimen treatments should be carefully documented in the Case Report Form (CRF). Any drugs affecting lower urinary tract function should also be recorded and statistically analyzed. Participants will receive instructions on health care education and lifestyle changes, including exercise, weight loss, smoking, etc.

### Acupuncture Group

The development of the acupuncture protocol was based on the consensus of acupuncturists from Guang’ anmen Hospital. The locations of the acupoints (Table 1) follow the World Health Organization (WHO) standard acupuncture point locations^[10]^. Bilateral Shenshu (BL23), Zhongliao (BL33), Huiyang (BL35), and Sanyinjiao (SP6)^[9]^ will be inserted using stainless steel disposable aseptic acupuncture needles (sizes 0.30×75 and 0.30×40 mm). Bilateral BL33 will be inserted to a depth of 50 to 60 mm with a 30° to 45° angle in an inferomedial direction using needles (0.30 mm in diameter, 75 mm in length). Bilateral BL35 will be inserted to a depth of 50 to 60 mm with a slightly superolateral direction using needles (0.30 mm in diameter, 75 mm in length). Bilateral BL23 and SP6 will be inserted vertically to a depth of 25 to 30 mm using needles (0.30 mm in diameter, 40 mm in length).

**Table 1.**
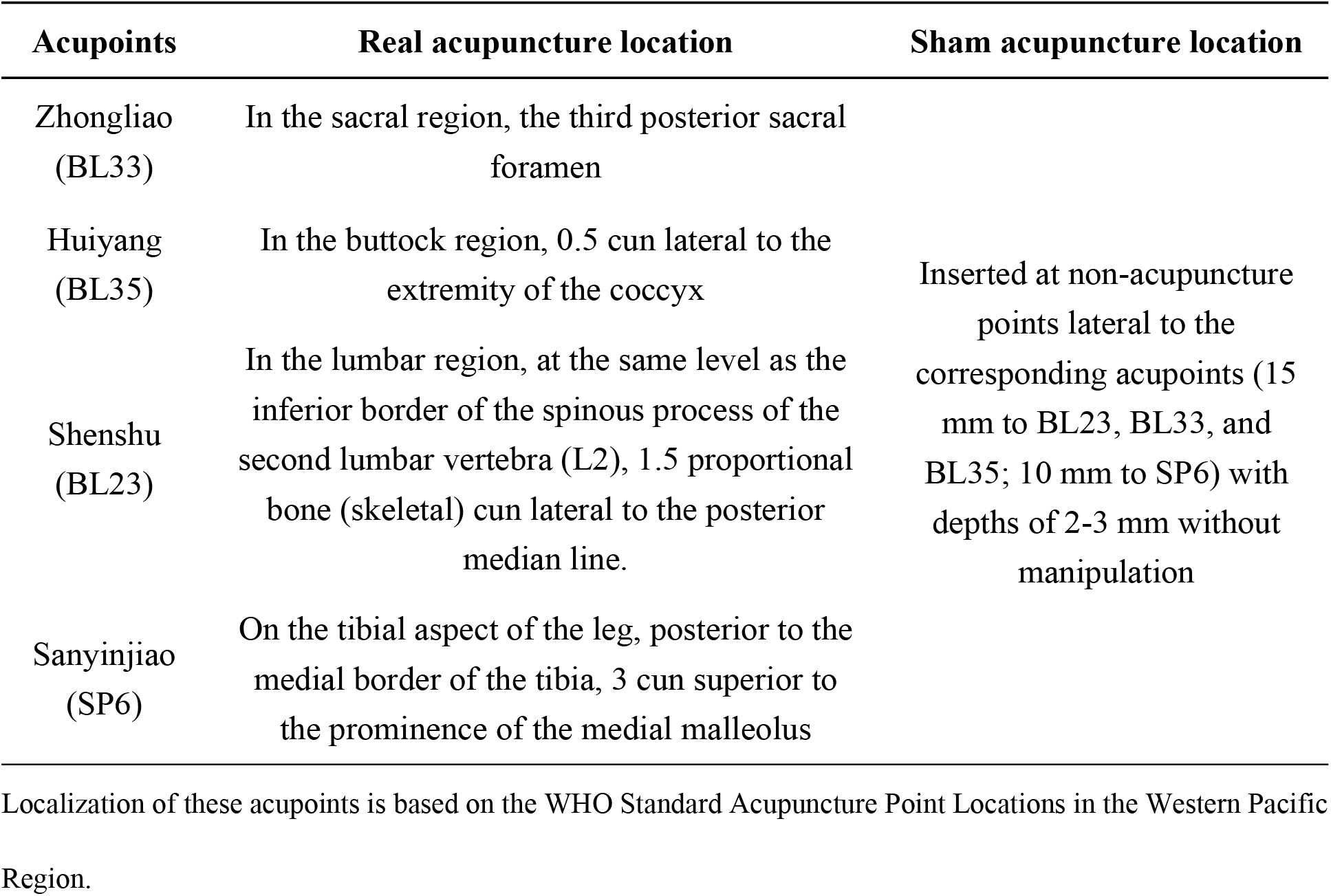
Locations of the selected acupoints for treating CP/CPPS

Manipulation of the needles by lifting and thrusting combined with twirling and rotating evenly will be performed until *deqi* occurs, a sensation of soreness, numbness, heaviness, and ache. Manipulations will be applied every 10 minutes, and each session will last for 30 minutes.

### Sham Acupuncture Group

Bilateral sham BL23, BL33, BL35 (15 mm to BL23, BL33, and BL35), and SP6 (10 mm to SP6) will be inserted by needles (size 0.2×25 mm) to a depth of 2 to 3 mm without manipulation and causing the feeling of *Deqi*.

### Outcome Measures

#### The Primary Outcome

Main outcome measures: after eight weeks of treatment and 24 weeks of follow-up, we will use the Chinese version of SF-36 (The MOS item short-form health survey) to evaluate the short-term effect of acupuncture for eight weeks to improve the QoL of patients with CP/CPPS, followed by 24 weeks of follow-up.

#### Secondary Outcomes

The secondary outcomes of this study include (a) a change in the International Prostate Symptom Score (IPSS). IPSS is a valid, reliable, and sensitive measure for patients with lower urinary tract symptoms (LUTS); it is widely used in clinical practice and research to determine the severity of LUTS, including incomplete bladder emptying, frequency of urination, intermittency, urgency, weak urine stream, straining and nocturia^[11,12]^; (b) change in the NIH-CPSI total score, the total score of the Chinese version NIH-CPSI. As a validated, self-reported questionnaire, NIH-CPSI is widely used to assess CP/CPPS symptoms^[13,14]^; (c) change in the NIH-CPSI subscales, including pain, urinary symptoms, and QoL impact. NIH-CPSI consists of 9 items divided into three discrete domains: pain (0-21 points), urinary symptoms (0-10 points), and QoL (0-12 points), with a total score of 0-43 points (a higher score indicates worse symptoms). A decline of at least 6 points in NIH-CPSI has been identified as the minimal clinically significant difference (MCID) ^[15]^ ; (d) changes in the visual analog scale (VAS) of pelvic floor pain. Secondary Outcomes will be assessed at the end of the 8^th^ and 24^th^ compared with baseline and the safety of acupuncture.

#### Incidence of Adverse Events

Patients will be asked to report any acupuncture-related adverse events (AEs) voluntarily. The researchers will record and evaluate. Acupuncture-related adverse reactions include bleeding, local infection, soreness, sweating, and so on.

#### Sample Size Estimation

Based on the previous pre-experimental study with a sample size of 20 people, 72% of the patients in the acupuncture group had a total NIH-CPSI score of 6 points or lower than the baseline at the 8^th^ week of treatment, and 36% of the patients were sham acupuncture. Take α=0.05, β=0.1, Power=0.9. 70 samples will be needed for 35 cases in each group.

#### Data Collection and Management

Collect data clearly and entirely in the CRF. All raw data sources will be retained, including CRFs, informed consent, and inspection results. An independent data administrator is responsible for completing the customer report format and will be required to double-enter the data for proofreading. All trial-related data will be kept for at least five years after publication. The reader can get the raw data by contacting the corresponding author. Patient information, including name, age, and phone number, will remain anonymous. An independent data and safety monitor board (DSMB) was established to ensure the integrity of the study data. The DSMB will review progress and decide whether premature closure is needed. Beijing Fengtai Hospital of Integrated Traditional and Western Medicine will designate at least three non-treatment practitioners to supervise the quality of the research

#### Quality control

All practitioners, including acupuncturists, research assistants, and statisticians, will be required to attend training to ensure the quality of this trial. The acupuncture operator must have the acupuncture doctor qualification certificate and independently undertake the clinical treatment for more than two years. Interventions will be carried out based on strict compliance with standardized operating procedures.

#### Statistical Analysis

Statistical analysis will be based on the principle of intention to treat, and third-party statisticians are unaware of group allocation and intervention procedures. The missing data will be estimated using the last observation carry-over method.

Normally distributed numerical data are presented as mean ± standard deviation, and non-normally distributed data are presented as medians with confidence intervals of 95%. For normally distributed data, repeated measures analysis of variance (ANOVA) was used for within-group and between-group comparisons to assess changes in continuous variables at different time points before and after the intervention; for non-normally distributed data, nonparametric tests were used to compare within groups and between groups. All data were analyzed using SPSS 22.0. All hypothesis tests were two-sided, and *p* values < 0.05 were considered statistically significant.

## Discussion

The incidence of CP/CPPS in men is high, and the cure rate is low. CP/CPPS is often associated with lower urinary tract dysfunction and negative emotion, cognition, and/or sexual dysfunction^[16]^. It seriously affects the QoL and mental health of the patients. In recent years, the pathogenesis of CP/CPPS has been the research focus. Many factors, such as urinary reflux^[17]^, pathogen infection^[18]^, inflammation, and oxidative stress^[19,20]^, may be involved in the pathogenesis of CP/CPPS. However, the existing treatment methods are not ideal because of their complicated clinical manifestations and lingering disease^[21]^. Some patients with prostate inflammation after the disappearance of pain still exist for a long time. Therefore, seeking an effective and safe non-drug therapy to reduce the urinary tract symptoms associated with patients, relieve pain, improve the QoL of patients with CP/CPPS, and reduce the side effects and medical costs of drug therapy is the key to treating CP/CPPS. Acupuncture has long been a non-pharmacological approach to pain treatment, with lasting effects of more than 12 months. A growing number of trials have shown that acupuncture may reduce symptoms compared with standard medical treatments^[22]^. In a Cochrane review of 20 non-pharmacological interventions evaluated for CP/CPPS^[23]^, only acupuncture and extracorporeal shock wave therapy likely result in symptom relief with reasonable safety. The study group found that 440 patients with CP/CPPS were randomly divided into an acupuncture group and a sham-acupuncture group. The two groups were treated with acupuncture and simulated acupuncture for eight weeks and 20 times, respectively. The results showed that the improvement of CP/CPPS symptoms in the acupuncture group was significantly better than in the control group. The difference between the two groups was clinically significant. The therapeutic effect could be maintained for half a year after stopping the treatment, and adverse events seldom occurred during the treatment. This study confirmed the long-term efficacy and safety of acupuncture in the treatment of CP/CPPS with high-quality clinical evidence.

Compared with the depth of the study on the pathogenesis of CP/CPPS by mainstream medicine in recent years, the study on the mechanism of the treatment of CP/CPPS by acupuncture and moxibustion is still in the initial stage. CP/CPPS is a kind of symptom-dependent disease. The most basic aim of treatment is to improve the symptoms. The current research results can not clarify the pathogenesis and pathway of the disease, so it is challenging to explore accurate treatment targets on this basis.

In recent years, with the continuous innovation of equipment and technology, neural regulation has been increasingly used in treating CP/CPPS. Acupuncture therapy can improve the symptoms by stimulating peripheral nerves, one of the modern nerve regulation models. For example, Zhongliao (BL33) is at the beginning of the gluteus maximus muscle, when the posterior branch of the lateral iliac artery and vein, and the point is at the beginning of the gluteus maximus muscle, the posterior branches of the second sacral nerve (S2) and the third sacral nerve (S3) pass through. The acupuncture sensation mostly stays in the local area when the superficial layer is punctured, which has a definite therapeutic effect on most of the local lumbosacral lesions. It will produce the needle sensation radiating to the anterior vulva, lower abdomen, and pelvic cavity, which has a better effect on the reproductive system, urinary system diseases, and regulating the function of the bladder when the acupuncture reaches a certain depth. Sanyinjiao (PC6) is a distal acupoint of the limbs; deep needling can stimulate the main trunk of the tibial nerve, which is a branch of the sciatic nerve and contains L4-S3 nerve fibers, stimulation of the tibial nerve can effectively stimulate the approach targeting the sacral plexus to improve the voiding symptoms in patients with CP/CPPS.

CP/CPPS is often associated with negative emotions, cognition, and/or sexual dysfunction and is a typical psychosomatic disorder. Acupuncture treatment can regulate peripheral nerves, inhibit pain signals from the pain conduction pathway to produce an analgesic effect, and improve pain threshold perception by improving pain-related emotion. But whether acupuncture can improve the QoL of patients with CP/CPPS is still uncertain. In this trial, we will use sham acupuncture to assess whether acupuncture improves the QoL and effectively relieves both short-and long-term symptoms in patients with CP/CPPS. With a 24-week follow-up of patients and strict quality control, our study will provide substantial evidence as to whether acupuncture can improve QoL in patients with CP/CPPS.

## Conclusions

The clinician not only needs to evaluate the disease comprehensively and consider the objective physiological indexes but also needs to understand the influence of the disease on the patient’s subjective feelings and functional status, as its physical, psychological, and social life and other aspects of the impact of patients to guide the rehabilitation and health decision-making. The current Randomized controlled trial protocols aim to investigate the efficacy and safety of acupuncture in treating patients with CP/CPPS. This trial is the first in the acupuncture field to use Randomized controlled trial QoL as a primary outcome measure.

## Data Availability

Deidentified research data will be made publicly available when the study is completed and published.

## Ethics Statement

The Ethics Committee of the Beijing Fengtai District Integrated Chinese and Western medicine hospital reviewed and approved these studies involving human participants.

Patients/participants provided their written informed consent to participate in the study.

## Author Contributions

The manuscript’s content is accurate and represents the authors’ work/opinions, not those of the sponsoring agent(s). Yu-long Ding put forward the idea of performing this review. Yu-long Ding wrote the initial paper. Yu-long Ding, Huai-Yu Wang, and Yuan-Ji revised and edited the manuscript. Shuo Zhang and Peng-Fei Yue drew the picture. Hong-Chao Zhao, Yan Guo, and Xiao-Di Xie summarized the tables. All authors have approved the submitted version. The authors declare that they have no conflict of interest.

## Funding

This work was supported by Fengtai District Health System Research Project (No. 2019-90)

## Conflict of Interest

The authors declare that the research was conducted without any commercial or financial relationships that could be construed as a potential conflict of interest.

## Acknowledgments

We want to thank professors Zhi-Shun Liu, Yu-Ying Cai, and Hong Zhao from Guang ‘anmen Hospital of China Academy of Chinese Medical Sciences for their guidance in the design and implementation of the project.

## Abbreviations

CP/CPPS: chronic prostatitis/chronic pelvic pain syndromes
QoL: quality of life
RCT: randomized controlled trial
NIH: National Institutes of Health
AF: atrial fibrillation
NSCLC: non-small cell lung cancer
Acu Group: acupuncture treatment group
Sham Group: sham needle control group
BUCM: Beijing University of Chinese Medicine
NIH-CPSI: National Institutes of Health Chronic Prostatitis Symptom Index
CRF: case report form
WHO: World Health Organization
BL: Bladder Meridian
SP: Spleen Meridian
SF-36: the MOS item short-form health survey
IPSS: international prostate symptom score
LUTS: lower urinary tract symptoms
MCID: minimal clinically important difference
VAS: visual analog scale
AEs: adverse events
DSMB: the data and safety monitor board
ANOVA: analysis of variance

## Notes

### Competing Interest Statement

The authors have declared no competing interest.

### Clinical Trial

ChiCTR2100051115

### Clinical Protocols

https://www.chictr.org.cn/hvshowproject.aspx?id=172850

### Funding Statement

This work was supported by Fengtai District Health System Research Project (No. 2019-90). Yu-Long Ding received this award. Webside: http://www.bjft.gov.cn/ftq/zfgg/201911/c8dba26474854711b3cfb4f0cfcef8c1.shtml

### Author Declarations

The Ethics Committee of the Beijing Fengtai District Integrated Chinese and Western medicine hospital reviewed and approved these studies involving human participants. Patients/participants provided their written informed consent to participate in the study.

